# SPEEDS: A Portable Serological Testing Platform for Rapid Electrochemical Detection of SARS-CoV-2 Antibodies

**DOI:** 10.1101/2021.05.16.21256907

**Authors:** Ran Peng, Yueyue Pan, Zhijie Li, Zhen Qin, James M. Rini, Xinyu Liu

**Author notes:** These authors contributed equally to this work (Ran Peng, Yueyue Pan).

## Abstract

The COVID-19 pandemic has resulted in a worldwide health crisis. Rapid diagnosis, new therapeutics and effective vaccines will all be required to stop the spread of COVID-19. Quantitative evaluation of serum antibody levels against the SARS-CoV-2 virus provides a means of monitoring a patient’s immune response to a natural viral infection or vaccination, as well as evidence of a prior infection. In this paper, a portable and low-cost electrochemical immunosensor is developed for the rapid and accurate quantification of SARS-CoV-2 serum antibodies. The immunosensor is capable of quantifying the concentrations of immunoglobulin G (IgG) and immunoglobulin M (IgM) antibodies against the SARS-CoV-2 spike protein in human serum. For IgG and IgM, it provides measurements in the range of 10.1 ng/mL − 60 µg/mL and 1.64 ng/mL − 50 µg/mL, respectively, and both antibodies can be assayed in 13 min. We also developed device stabilization and storage strategies to achieve stable performance of the immunosensor within 24-week storage at room temperature. We evaluated the performance of the immunosensor using COVID-19 patient serum samples collected at different time points after symptom onset. The rapid and sensitive detection of IgG and IgM provided by our immunosensor fulfills the need of rapid COVID-19 serology testing for both point-of-care diagnosis and population immunity screening.

## 1. Introduction

Since the onset of the coronavirus disease (COVID-19) pandemic in March 2020, this global health crisis has caused 146,054,107 infections and 3,092,410 fatalities (as of April 25^th^, 2021) (World Health Organization, 2021). To mitigate the impact of the pandemic, effective prevention of the spread of COVID-19 is urgently needed. It has been demonstrated in many countries that the COVID-19 pandemic could be controlled with a series of measures, such as rapid diagnostics, infection and contact tracing, large-scale vaccination, and immunotherapy (Bhalla et al., 2020; Carter et al., 2020; Cheng et al., 2020a; Ji et al., 2020; Jin et al., 2020; Qin et al., 2020; Ravi et al., 2020; Udugama et al., 2020). Serological assays for determining antibody responses against the severe acute respiratory syndrome coronavirus 2 (SARS-CoV-2) provide ammunition for these pandemic control tactics. The role of serological testing in clinical diagnostics and public health measures has been debated ever since the beginning of the pandemic (Tré-Hardy et al., 2020). For instance, it has been argued that serological testing could serve as an alternative diagnostic method in countries and regions with limited access to molecular testing (Peeling et al., 2020). It can also be used as a complement to a polymerase chain reaction (PCR)-based diagnosis (Udugama et al., 2020). Another widely recognized use of serological testing is to determine the past infection history of individuals, allowing for longitudinal immunity tracking.

As the gold-standard diagnostic method for COVID-19, reverse-transcription PCR (RT-PCR) detects conserved regions of the SARS-CoV-2 RNA genome. However, it has a relatively high cost per test and requires costly equipment and highly-trained laboratory personnel (Chaimayo et al., 2020; Corman et al., 2020; Moore et al., 2020). This makes the RT-PCR test less accessible in many developing countries (Giri and Rana, 2020; Mannan and Nseluka, 2020). Other diagnostic methods have been developed as alternatives to RT-PCR, including rapid viral antigen testing, loop-mediated isothermal amplification (LAMP)-based rapid RNA testing, and serological testing (Ahmadivand et al., 2021; Fabiani et al., 2021; Lee et al., 2021; Liu et al., 2021; Raziq et al., 2021; Torrente-Rodríguez et al., 2020; Vabret et al., 2020; Yousefi et al., 2021). Because of its short turnaround time and low cost, serological testing has been recommended as an effective method for COVID-19 patient diagnosis, especially in countries/regions with limited capacity for large-scale molecular testing (Peeling et al., 2020). In addition, serological testing can also be used for population screening and contact tracing (Mathur and Mathur, 2020), as well as long-term population surveillance that could provide a reference for setting/adjusting pandemic control measures (Winter and Hegde, 2020). Since antibody levels can persist for months after a SARS-CoV-2 infection, serological testing is also suitable for longitudinal immunity assessment (Isho et al., 2020; Yongchen et al., 2020).

A variety of immunoassay platforms have been developed, by both academic laboratories and industrial companies, for the detection of SARS-CoV-2 antibodies. A widely adopted commercial immunoassay platform for COVID-19 serological testing is the lateral flow test (LFT) strip. Many LFT products have received regulatory approvals in different countries for COVID-19 diagnosis. Despite their ease of operation, short assay time, and low cost, these strips usually provide relatively low clinical sensitivity and specificity (Wu et al., 2020; C. Zhang et al., 2021). The conventional laboratory enzyme-linked immunosorbent assay (ELISA) is so far the most promising serology testing method for COVID-19 diagnosis because of its high sensitivity and specificity (Adams et al., 2020; GeurtsvanKessel et al., 2020). However, it requires laboratory infrastructure and equipment and takes hours for a single run (Kasetsirikul et al., 2020b; Roy et al., 2020; Tan et al., 2020a). Targeting rapid and sensitive COVID-19 serology testing at the point of care (POC), a variety of portable immunosensing platforms have been developed based on microfluidics and biosensor technologies (Fabiani et al., 2021; Kim et al., 2021a; Lee et al., 2021; Li et al., 2021; Raziq et al., 2021; Roda et al., 2021; Tan et al., 2020a; Torrente-Rodríguez et al., 2020; Yakoh et al., 2021; Zakashansky et al., 2021; C. Zhang et al., 2021; Z. Zhang et al., 2021; Zhao et al., 2021).

In this paper, we present a portable <u>S</u>erological testing <u>P</u>latform for rapid <u>E</u>lectroch<u>E</u>mical <u>D</u>etection of <u>S</u>ARS-CoV-2 antibodies (SPEEDS). It is based on a low-cost electrochemical immunosensor that uses ELISA to quantitate immunoglobulin G (IgG) or immunoglobulin M (IgM) antibodies against the SARS-CoV-2 spike protein (S-protein) found in serum. The SPEEDS platform only takes 13 minutes for a complete assay and the electrochemical immunosensor can be batch-fabricated at low cost. Through simple device packaging, the prepared ready-to-use immunosensor chips can be stored at room temperature without performance deterioration for at least 24 weeks. We achieved wide measurement ranges of 10.1 ng/mL − 60 µg/mL and 1.64 ng/mL − 50 µg/mL for human monoclonal anti-SARS-CoV-2 IgG and IgM, respectively, which cover typical antibody levels in convalescent sera, as well as the sera of patients with both mild and severe COVID-19 infections. Using the SPEEDS platform, we performed serological testing of 30 patient samples and demonstrated satisfactory clinical performance. The SPEEDS platform provides a low-cost and reliable diagnostic tool for POC serological testing of COVID-19.

## 2. Materials and methods

### 2.1 Materials and reagents

Streptavidin (N7021S) was purchased from New England Biolab (Whitby, ON, Canada). The blocking reagent for ELISA (11112589001), immunoassay stabilizer (s0950), alkaline phosphatase (ALP) conjugate stabilizer (76696), biotin (5-fluorescein) conjugate (53608), human serum (P2918), 10× Tris-acetate-EDTA (TAE) buffer (T9650) used to prepare 1× TAE buffer, K_3_[Fe(CN)_6_] and sulfuric acid (258105) were purchased from Sigma-Aldrich (Oakville, ON, Canada). ALP-conjugated goat anti-human IgG (ab97222) and goat anti-human IgM (ab97202) were purchased from Abcam (Toronto, ON, Canada). The electrochemical substrate P-aminophenyl phosphate (A-292-500) was purchased from Gold Biotechnology (St. Louis, MO, USA). Phosphate buffered saline (PBS) (10010023) and distilled water (15230162) were purchased from Thermo Fisher Scientific (Ottawa, ON, Canada). The ELISA diluent (3652-D2) was purchased from Mabtech (Cincinnati, OH, USA). The CR3022 IgG and IgM antibodies were constructed by cloning the CR3022 Fab into a human IgG1 and human IgM framework, respectively (Ter Meulen et al., 2006). Biotinylated SARS-CoV-2 S-protein receptor-binding domain (RBD), human monoclonal anti-SARS-CoV-2 CR3022 IgG and IgM were produced as previously described (Abe et al., 2020). Human total IgG (I4506) was purchase from Sigma-Aldrich. Patient serum samples (CoV-PosSet-S1) were purchased from RayBiotech (Peachtree Corners, GA, USA). The patient sample testing was approved by the research ethics boards at the University of Toronto (protocol number: 40357). Carbon ink (E3456) and Ag/AgCl ink (E2414-250G) for screen printing were purchased from Ercon Inc. (Wareham, MA, USA), and PDMS elastomer (SYLGARD™ 184) for hydrophobic line printing was purchase from Dow Corning (Midland, MI, USA). Blotting paper (Whatman Blotting Membranes, catalog #: 3030-6185, Cytiva) was used during the operation of fluids. Chip substrates, including Whatman Blotting Membranes (WHA3001861) polyethylene terephthalate (PET) transparent film (CG7060), were purchased from Sigma-Aldrich and Amazon, respectively. Wax printing (Xerox 8560DX) was applied to pattern hydrophobic zones on the sensor substrates. The protection film (3082307) for screen-printing was purchased from Dollarama, and airtight bags (B07QCM4MZ8) and desiccant (B00E880DYS) for device storage were purchased from Amazon.

### 2.2 Design of the SPEEDS platform

The SPEEDS platform, as shown in **Figure 1a**, consists of a custom-made electrochemical immunosensor and a commercial handheld potentiostat (EmStat3 Blue, PalmSens). The immunosensor includes three screen-printed electrodes (a carbon working electrode, a carbon counter electrode, and an Ag/AgCl reference electrode) on a PET transparent film, forming a three-electrode electrochemical cell. It can be directly inserted into a chip slot of the handheld potentiostat for electrochemical signal readout, and the testing data are transmitted, through a Bluetooth connection, to a smartphone (HUAWEI Mate20 X). The SPEEDS platform can address the urgent need for rapid COVID-19 serological testing in many settings including airports, customs/borders, long-term home cares, schools, and densely populated workplaces. By providing rapid and sensitive quantification of SARS-CoV-2 antibodies in serum samples, it can complement the PCR-based laboratory test and the rapid antigen COVID-19 test. It also allows for a retrospective study of COVID-19 infection via population immunity screening and tracking (**Figure 1a**).

**Figure 1.**
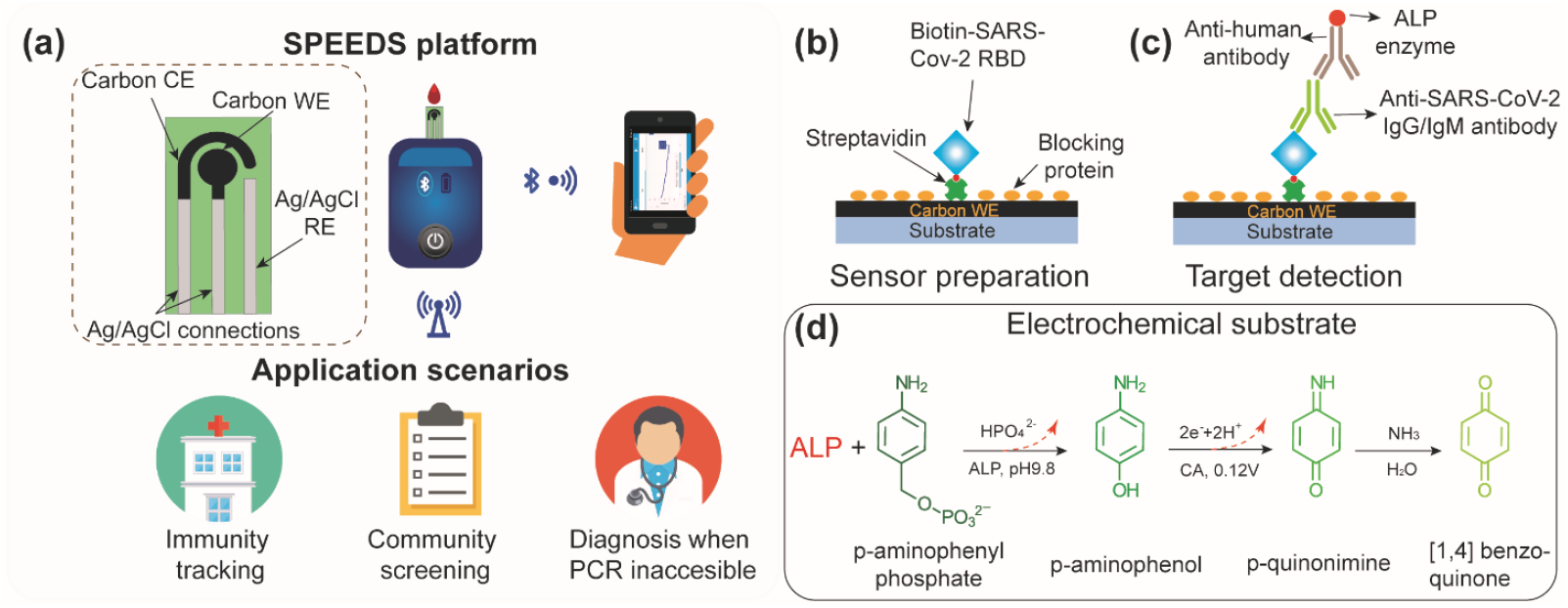
Design of the SPEEDS platform for detection of IgG and IgM antibodies against SARS-CoV-2 spike protein in human serum. (a) Schematic illustration of the SPEEDS platform and its application scenarios. The platform includes an electrochemical immunosensor and a handheld potentiostat. CE: counter electrode, WE: working electrode, and RE: reference electrode. The potentiostat can transmit testing results to a cell phone. (b) Schematic of surface functionalization of the WE with biotinylated RBD protein as the capture probe. (c) Schematic of capturing anti-SARS-CoV-2 IgG or IgM on the WE and subsequently labelling it with ALP-conjugated detection antibody. (d) Oxidation of the electrochemical substrate (pAPP) during chronoamperometry (CA) and production of the CA current.

The detection of the SARS-CoV-2 antibodies on the immunosensor is based on an electrochemical ELISA, as illustrated in **Figure 1b-c**. Streptavidin is first immobilized, through physical absorption, on the working electrode (WE) of the immunosensor followed by the immobilization of the capture probe via streptavidin/biotin binding (**Figure 1b**). The biotinylated SARS-CoV-2 spike receptor-binding domain (RBD) protein is used as the capture probe that provides high specificity to SARS-CoV-2 IgG and IgM antibodies. During each test, the SARS-CoV-2 IgG or IgM antibodies in the sample are captured on the WE by the RBD capture probe (**Figure 1c**). Then, alkaline phosphatase (ALP)-labeled anti-human detection antibody, specifically against IgG or IgM antibodies, is added to bind to the captured IgG or IgM antibody. Lastly, the electrochemical substrate of ALP, p-aminophenyl phosphate (pAPP), is added to the immunosensor to react with the ALP for chronoamperometric (CA) measurement (**Figure 1d**). A higher concentration of the IgG or IgM antibody leads to a higher density of the immobilized ALP on the WE and thus a higher CA current. Therefore, the concentration of SARS-CoV-2 IgG or IgM antibody can be quantified based on the CA current.

### 2.3 Fabrication and preparation of the immunosensor

#### 2.3.1 Fabrication of the immunosensor

The electrochemical immunosensor, as shown in **Figure 1a** and **S1a**, contains three electrodes (carbon WE, carbon CE, and Ag/AgCl RE) screen-printed on a PET substrate, and all three electrodes are confined in a hydrophilic reaction zone patterned through solid wax printing. The screen-printing protocol for patterning carbon and Ag/AgCl electrodes has been reported previously (Zhao and Liu, 2016). Briefly, a hydrophilic reaction zone was first patterned by wax printing on a PET substrate, and then the PET substrate was heated at 100°C for 2 min. Then, the Ag/AgCl RE and the ‘tail’ connections of WE and RE were screen-printed manually on the substrate using a stencil film with the electrode shapes cut by a blade cutter (CM350, Brother ScanNCut2 Cutting Machine). Finally, the carbon WE and CE were screen-printed. For drying the printed electrodes, the PET substrate was baked at 50 °C for 60 min after each round of screen printing. In a single fabrication procedure, we batch-fabricated immunosensors on four letter-sized PET films (each contains 96 immunosensors, see **Figure S1b**), and the entire fabrication process (for four PET films containing 384 immunosensors) typically took 150 minutes.

To prevent reagent solutions added to the WE from wicking out of the circular WE reaction zone through the WE tail connection (**Figure 2a**), a thin PDMS barrier was printed onto the WE top surface at the interconnection of the circular WE reaction zone and the tail connection through contact printing (**Figure S1a**). To print the PDMS barrier, a fishing line of 100 µm in diameter coated with PDMS precursor (w/w ratio of the base and curing agent: 10:1) was brought into contact with the WE top surfaces of eight curved immunosensors attached on a 3D-printed bending mold (10 cm in diameter), as shown in **Figure S1a**. The PDMS barrier can efficiently confine solutions added to the reaction zone of a WE, leading to consistent performance of each immunosensor. Surface functionalization and stabilization of capture proteins on the WE (see the next section for protocols) were conducted on the bare immunosensor chips to allow electrochemical ELISA of SARS-CoV-2 antibodies. The chips were then dried in air and stored in nitrogen-filled airtight bags with desiccant for long-term storage (see **Figure S2a**). The ready-to-use immunosensors can be stored at room temperature.

**Figure 2.**
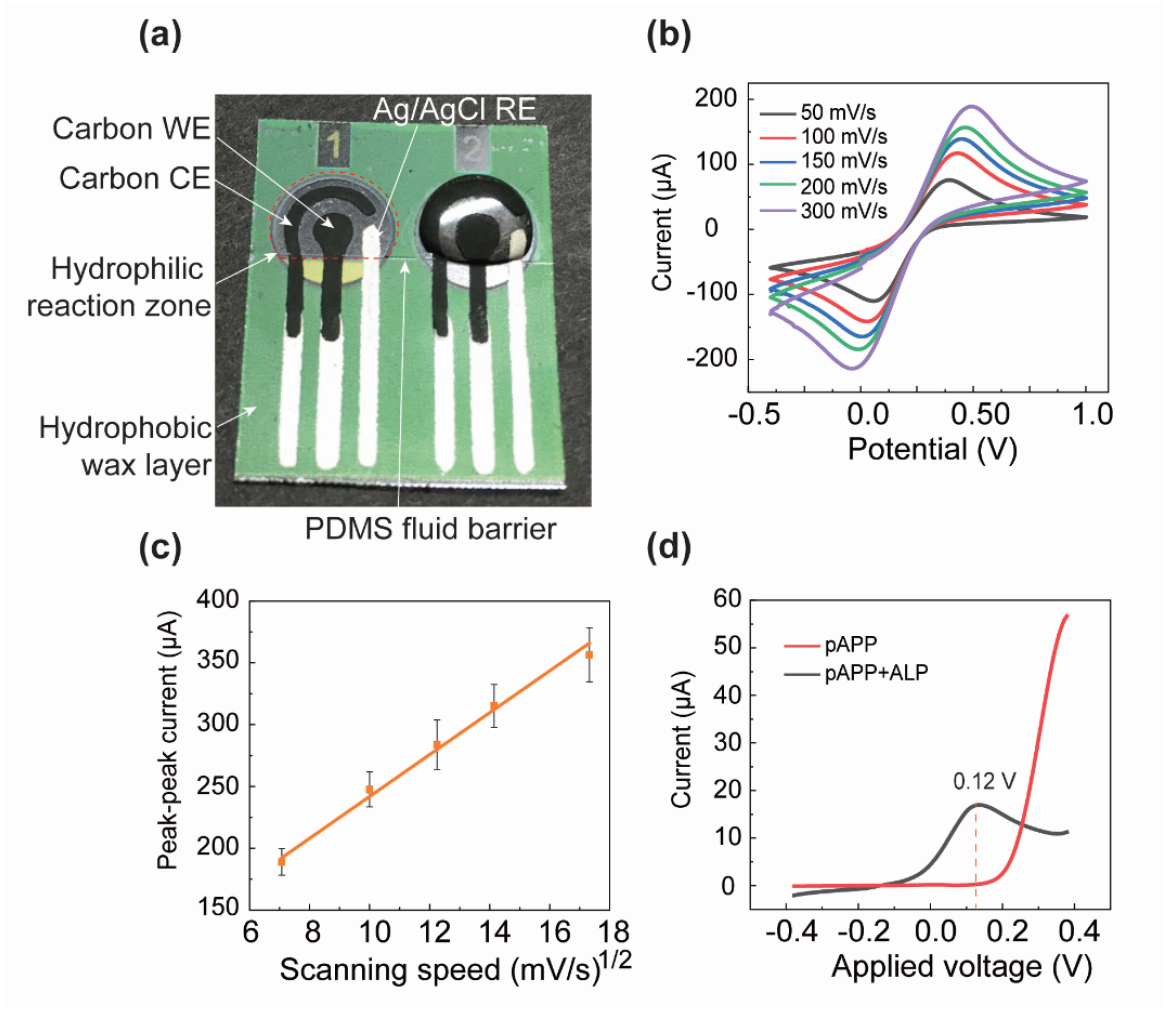
Fabrication and characterization of the electrochemical immunosensor. (a) A photograph of the screen-printed three-electrode immunosensor loaded with sample solution in the reaction zone. The reaction zone is defined by a wax-printed cycle and a thin PDMS line barrier printed at the “tail” of the electrode. (b) Measurement of cyclic voltammetry (CV) curves at scan rates of 50 mV/s, 100 mV/s, 150 mV/s, 200 mV/s, 300 mV/s. (c) The peak-peak current of the CV curve versus the square root of the scan rate (n=5). (d) The oxidation current-voltage signals measured from electrochemical immunosensors loaded with the electrochemical substrate pAPP and a mixture of pAPP and ALP-conjugated detection antibody, under the condition of hydrodynamic linear sweeping voltage in the range of −0.4 V and 0.4 V with a scanning speed of 100 mV/s.

#### 2.3.2 Activation, biofunctionalization, stabilization and long-term storage of the immunosensor

Before surface functionalization of the WE with capture probes, the reaction zone of the WE was loaded with 100 µL of 0.18 M sulfuric acid (diluted in 1× PBS) and then activated for 3 min with a 1.3 V anodic voltage (against to RE). This activation process creates abundant active carboxylic groups on the WE carbon surface and thus improves the sensitivity of the device (Díaz-González et al., 2005). After washing with DI water and then 1× PBS, the WE underwent a series of surface functionalization steps including i) immobilization of streptavidin (SA), ii) hybridization of biotinylated RBD, and iii) blocking of the void locations of the WE with blocking proteins (**Figure 1b**). Finally, the biofunctionalized WE surface was then stabilized with immunoassay stabilizer. Unless otherwise stated, all the washing steps in the WE biofunctionalization process are finished by dispensing 50 µL buffer (1× PBS or DI water) to the reaction zone, removing the solution by pipetting, and absorbing the residual buffer with blotting paper.

The immobilization of the RBD capture probe on the WE relies on the widely adopted biotin-streptavidin (SA) interaction (Diamandis and Christopoulos, 1991; Feng et al., 2021). First, the SA was immobilized on the WE by physical absorption. For this step, we investigated the absorption efficiency of SA on the WE with different incubation durations and studied the effect of washing cycles on the stability of SA immobilization. In our experiments, 5 µL of 1 mg/mL SA (in 1× PBS) was added on the WE reaction zone and incubated at 4 °C for different durations in the range of 1∼24 h. After incubation, the residual SA was washed off the WE by PBS, and 5 µL of 20 µM biotin (5-fluorescein) conjugate was added on the WE and incubated for 10 min to examine the efficiency of SA immobilization. All the WE surfaces were imaged under a fluorescence microscope (under 20× objective), and the average fluorescence intensities of the WE surfaces were analyzed in ImageJ.

**Figure S3b-f** show typical fluorescent images of the SA-immobilized WE surfaces after 1, 4, 8, 12 and 24 h incubation in the SA solution and 3 times of PBS washing after SA incubation, respectively. One can observe uniform SA coating on the WE surface for all five SA incubation durations. **Figure S3g** illustrates the average fluorescence intensity of the WE surface as a function of the incubation time. One can see there is an obvious increasing trend of the amount of immobilized SA with the incubation time, indicating longer incubation time improves the immobilization efficiency. To investigate the stability of the SA immobilization on the WE, the fluorescence intensities of the WE surfaces, which were modified with 12 h SA incubation and then washed by 50 µL of PBS two, three or five times, were analyzed. The results (**Figure S3h**) indicate that the average fluorescence intensity of the WE surface after five times of PBS washing is only 5.6% lower than that of the WE after twice of PBS washing, suggesting highly stable immobilization of SA on the WE through physical absorption. Based on the above data, the modification condition of 24-hour SA absorption and three times of PBS washing were adopted for preparing all the immunosensors for the following experiments.

After the SA immobilization, the immunosensors were grafted with SARS-CoV-2 spike RBD protein. 5 µL of biotinylated RBD (50 µg/mL in PBS, unless otherwise stated) was pipetted onto the SA-immobilized reaction zone of the WE and incubated for 20 min. The WE surface was then washed by PBS three times. After that, 30 µL of commercial blocking reagent was added onto the WE reaction zone and incubated for 30 min. The WE reaction zone was washed again by PBS three times. Finally, 30 µL of commercial immunoassay stabilizer was added onto the WE reaction zone and incubated for another 30 min to stabilize the immobilized capture proteins. The residual stabilizer solution was removed with a pipette and the device was dried in air, leaving a thin protective stabilizer layer on the WE reaction zone. All the incubation and drying steps were performed in air at room temperature (21°C). The dried immunosensors were stored in a nitrogen-filled airtight bag with desiccant for long-term storage. The total material cost of an immunosensor chip is USD 2.10 in small quantity (see itemized material costs in **Table S1**).

### 2.4 Serological test procedure

For each serological test, the ready-to-use immunosensor was initially activated by adding 30 µL of immunoassay stabilizer to the WE reaction zone for one-minute incubation followed by washing with 50 µL of PBS twice. The immunosensor tests serum samples with five times dilution (in ELISA diluent), which reduces the interference from serum proteins. 5 µL of diluted serum sample was pipetted on a freshly reactivated WE and incubated for 1 min to enable the capture of the target antibody on the WE, followed by washing with PBS three times. Then, 5 µL of ALP-labeled anti-human antibody (20 µg/mL, diluted in the ALP conjugate stabilizer) was added to the WE and incubated for 3 min, which was followed by another three times of PBS washing. The ALP-antibody catalyzes the oxi-reductive reaction of pAPP. As the last step, 40 µL of pAPP (6 mM, diluted in 1× TAE buffer) was added on the WE and incubated for 5 min to reach reaction equilibrium. CA measurement was finally conducted and the stabilized faradaic current was measured 2 minutes after the stepwise CA potential was applied. For each test, the whole process took approximately 13 min.

### 2.5 Data acquisition and statistical analysis

The CA measurement was performed by using a portable potentiostat (EmStat3 Blue, PalmSens) capable of Bluetooth communication with a smartphone (HUAWEI Mate20 X). The device calibration data were fitted into sigmoidal curves using the Boltzmann equation 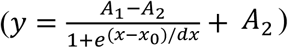 and the coefficient of determination (COD) *R*^*2*^ was calculated (see **Table S2** for fitting parameters). The limit of detection (LOD) of the immunosensor was calculated to be the antibody concentration corresponding to three times the standard deviation above the average output signal at zero concentration (Shrivastava and Gupta, 2011). Two-sample *t*-testing was used to analyze the cross-reactivity and long-term storage data with a confidence level of 95%. All statistical analyses were conducted in OriginPro 2018 (OriginLab).

### 2.6 Patient sample testing

A cohort of 20 COVID-19 positive serum samples (confirmed by RT-PCR) and 10 COVID-19 negative serum samples (collected before the pandemic) were tested using our immunosensors. Before the patient sample testing experiments, all the serum samples were treated with 0.5% Triton X-100 for 10 minutes to inactivate any potential infectious viruses. For each serum sample, both IgG and IgM tests were repeated twice each. All the numerical data are listed in **Table S3**.

## 3. Results and discussion

### 3.1 Electrochemical characterization of the immunosensor

To verify the electrochemical performance of the three-electrode immunosensor, 40 µL of 10 mM K_3_[Fe(CN)_6_] (diluted in 1 M KCl) was added to the WE of an immunosensor, and cyclic voltammetry (CV) measurements were conducted at different scan rates. As shown in **Figure 2b**, the measured cyclic voltammograms reveal typical reversible electrochemical reactions at all scan rates. The peak-peak current of the cyclic voltammogram is linearly proportional to the square root of the scan rate, further confirming that the immunosensor is a reversible electrochemical system. For testing serum samples, we chose CA measurement for its lower signal-to-noise ratio and thus higher sensitivity over CV measurement (Dungchai et al., 2009; Zhao et al., 2013). To determine the optical oxidation voltage of pAPP for CA measurement, hydrodynamic linear sweeping was performed on pure pAPP solution (6 mM in 1× TAE buffer) and pAPP plus ALP-labeled detection antibody solution (20 µg/mL in ALP conjugate stabilizer) at a 100 mV/s scan rate in the range of −0.4 V and 0.4 V. As shown in **Figure 2d**, the highest oxidation peak was obtained at 0.12 V. Therefore, 0.12 V was adopted as the excitation voltage for CA testing of the serum samples.

### 3.2 Calibration of the immunosensor

For device calibration, CR3022 IgG or IgM (concentration range: 500 pg/mL to 60 μg/mL) spiked in five-times diluted human serum were tested. **Figure 3a** and **3b** show the typical CA current curves measured from IgG-spiked and IgM-spiked serum samples at different concentrations. To minimize errors caused by non-Faradaic current, the steady-state current value at ∼120 s after the excitation voltage step was collected as the immunosensor output (zoomed-in views of the CA curves shown in **Figure 3c** and **3d**). Using IgG as the model analyte, we also investigated the effect of biotinylated RBD capture protein concentration (in the range of 10-100 µg/mL) on the performance of the immunosensor. **Figure S4** shows the calibration curves of IgG detection with different RBD concentrations (concentrations of all other reagents are the same as described in Section 2.3), suggesting that low concentrations of RBD (10 and 25 µg/mL) would result in unsatisfactory sensitivity when quantifying the target antibody at low concentrations while a high RBD concentration (100 µg/mL) improves the testing sensitivity but also raises the reagent cost. Considering the assay sensitivity and the manufacturing cost, 50 μg/mL biotinylated RBD was adopted for final device calibration experiments. **Figure 3e** and **3f** illustrate the calibration curves for the detection of CR3022 IgG and IgM in serum, respectively. The LOD values of the immunosensor for IgG and IgM are 10.1 ng/mL and 1.64 ng/mL, respectively.

**Figure 3.**
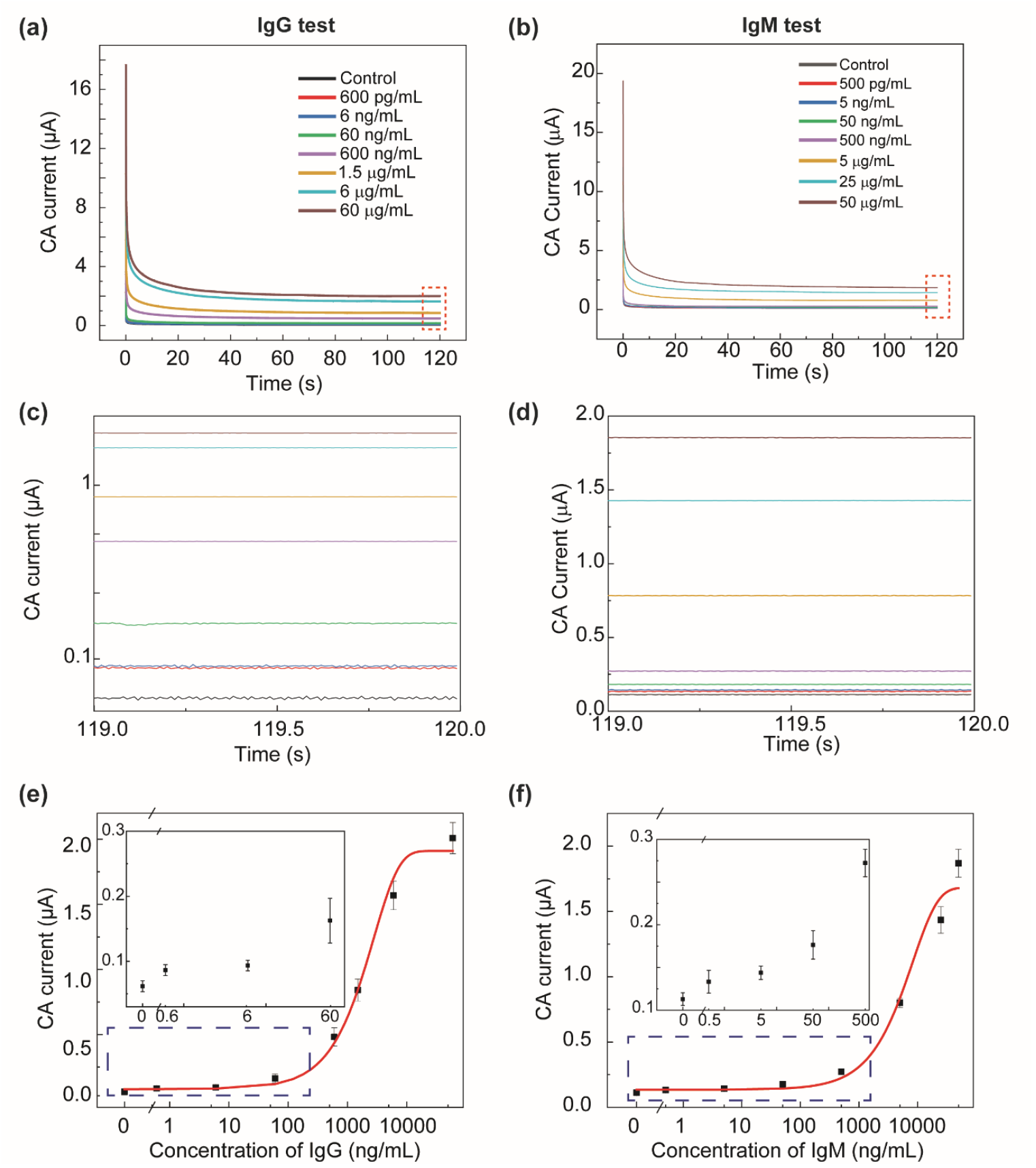
Calibration of the immunosensor for detecting anti-SARS-CoV-2 IgG and IgM in five-fold-diluted serum samples. (a)(b) Representative CA current curves measured on five-fold-diluted serum spiked with the (a) CR3022 IgG antibody and (b) CR3022 IgM antibody at different concentrations. The quasi-steady-state current at 120 s was used as the signal readout for device calibration. (c)(d) The zoomed-in views of the CA current curves in (a)(b) during 119-120 s, respectively. (e) Calibration curve of the immunosensor for IgG detection at 600 pg/mL, 6 ng/mL, 60 ng/mL, 600 ng/mL, 1.5 μg/mL, 6 μg/mL and 60 μg/mL. (f) Calibration curve of the immunosensor for IgM detection at 500 pg/mL, 5 ng/mL, 50 ng/mL, 500 ng/mL, 5 μg/mL, 25 μg/mL, and 50 μg/mL. For both IgG and IgM calibration, n=7 for current data at non-zero concentrations and n=10 for current data (background signals) at the zero concentration.

The IgG concentration in different individuals during and after SARS-CoV-2 infection varies significantly. Fortunately, there have been many studies revealing that the peak concentrations of antibodies against the SARS-CoV-2 RBD in COVID-19 patient serum are typically in the 10’s of µg/mL, with only a few cases reaching 100 µg/mL (Ibarrondo et al., 2020; Iyer et al., 2020; Ma et al., 2020; Terpos et al., 2020; Torrente-Rodríguez et al., 2020). Therefore, we chose to set the measurement amplitude of our immunosensor to be up to 60 µg/mL. Monoclonal antibody CR3022 was chosen in device calibration for its ability to bind the SARS-CoV-2 S-protein RBD. The calibration curves show the typical sigmoidal relationship between the CA current and the antibody concentration in the measurement range. Owing to the stable complex of the capture probe and the target antibody, and the strong reaction realized in our finalized assay conditions, wide detection ranges have been achieved for both IgG (10.1 ng/mL – 60 μg/mL) and IgM (1.64 ng/mL to 50 μg/mL) antibodies. As mentioned above, the antibody concentrations among patients differ significantly, emphasizing the need for a wide measurement range. Our device responds to both antibody isotypes over wide ranges with LODs at the ng/mL level, making it suitable for COVID-19 early diagnosis and longitudinal quantification of SARS-CoV-2 antibodies.

### 3.3 Cross-reactivity testing

The cross-reactivity of the immunosensor was also investigated. Since IgG is the most abundant antibody isotype in blood and persists for months while IgM is induced earlier and decays rapidly, we mainly focused on the cross-reactivity between anti-SARS-CoV-2 IgG and total human IgG. We used total human IgG produced before the COVID-19 outbreak as the interference antibody. Experiments were conducted with antibodies spiked in five-fold-diluted human serum (see results in **Figure 4a**): i) CR3022 IgG (600 ng/mL), ii) CR3022 IgG (600 ng/mL) and total human IgG (2.5 µg/mL), and iii) total human IgG (2.5 µg/mL). In all cases, ALP-labeled anti-human IgG antibody was used as the detection antibody. All experiments were repeated 5 times on different immunosensors. It should be noted that the incubation time was deliberately increased to 5 min to further amplify signals from any possible cross-reactivity. For the serum samples (**Figure 4a**), the average CA current of the “CR3022 IgG + total human IgG” sample (0.499 µA) was similar to that of the pure “CR3022 IgG” sample (0.493 µA), but both types of sample yielded average output currents much larger than the pure “total human IgG” sample (0.0962 µA). Experiments with antibody-spiked PBS generated consistent results (**Figure S5a**). These results show that our immunosensor for CR3022 IgG detection has negligible cross-reaction with non-specific total human IgG.

**Figure 4.**
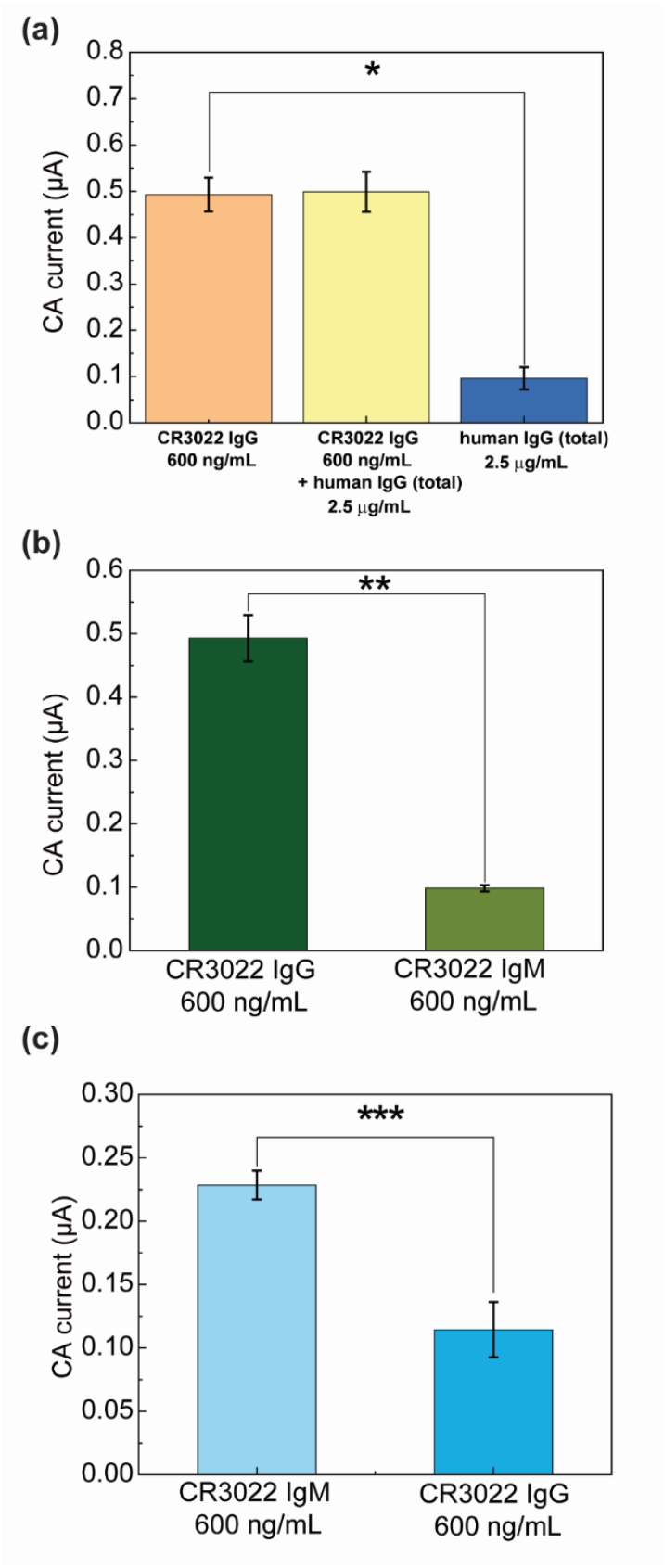
The cross-reactivity testing of the immunosensor for detection of anti-SARS-CoV-2 IgG and IgM. (a) The cross-reactivity data of CR3022 IgG detection using total human IgG antibody as the interference protein. *p=3.58×10^−8^. (b) The cross-reactivity data of CR3022 IgG detection using CR3022 IgM as the interference protein. **p=1.01×10^−8^. (c) The cross-reactivity data of CR3022 IgM detection using CR3022 IgG as the interference protein. ***p=6.27×10^−6^. In (a-b), ALP-labelled anti-human IgG was used as the detection antibody. In (c), ALP-labelled anti-human IgM was used as the detection antibody. For all the testing data, n=5.

The cross-reactivity between CR3022 IgG and IgM was also tested on our device in both diluted human serum (**Figure 4b-c)** and PBS **(Figure S5)**. In the IgG assay, ALP-labelled anti-human IgG was used as the detection antibody, CR3022 IgG (600 ng/mL) and CR3022 IgM (600 ng/mL) were spiked in five-fold-diluted human serum and detected separately (mean currents were 0.493 µA and 0.0984 µA, respectively; see results in **Figure 4b**). In the IgM assay, ALP-anti-human IgM was used as the detection antibody, CR3022 IgM (600 ng/mL) and CR3022 IgG (600 ng/mL) were spiked in diluted human serum and detected separately (mean currents were 0.228 µA and 0.114 µA respectively; see results in **Figure 4c**). All experiments were repeated five times on different immunosensors. From **Figure 4b** and **4c**, one can observe that the cross-reactivity between CR3022 IgG and IgM testing on our immunosensor is not significant (p-values: 1.01×10^−8^ and 6.27×10^−6^, respectively). The negligible cross-reactivity is due to the proper assay design as well as the high specificity of the secondary antibodies specific for IgG or IgM. It has been proven that S-protein-based serology tests showed less cross-reactivity than nucleocapsid protein (N-protein)-based assays (Amanat et al., 2020; Cheng et al., 2020b; Okba et al., 2020).

### 3.4 Long-term storage testing

The device stability over long-term storage is an important factor in practical use of our immunosensor. The possible degradation of the capture proteins on the immunosensor could result in testing performance decline. The performance stability of our immunosensors was investigated throughout a 24-week storage period. All the immunosensors (with immunoassay stabilizers) were prepared in the same batch and packed in the nitrogen-filled airtight bags with desiccant, in either laboratory environment (21°C and 30-50% humidity) or a refrigerator (4 °C). CR3022 IgG diluted in PBS at 6 µg/mL was used for testing the performance stability of the immunosensor.

The testing data (**Figure 5**) show that, for both room temperature and refrigerated storage conditions, no significant variation in the immunosensor output was observed over 24 weeks. These results confirm the high storage stability and performance reproducibility of our laboratory-made immunosensor, further demonstrating the high feasibility of our platform for practical COVID-19 serology testing. Among the recently published reports on COVID-19 serological biosensors (Kasetsirikul et al., 2020b; Tan et al., 2020b; Torrente-Rodríguez et al., 2020), limited long-term storage studies have been conducted. Our immunosensor shows a comparable shelf life to existing commercial COVID-19 serology tests such as the Abbott BinaxNOW Ag Card Tests (6 months) and cPass™ SARS-CoV-2 Neutralization Antibody Detection Kit (6 months).

**Figure 5.**
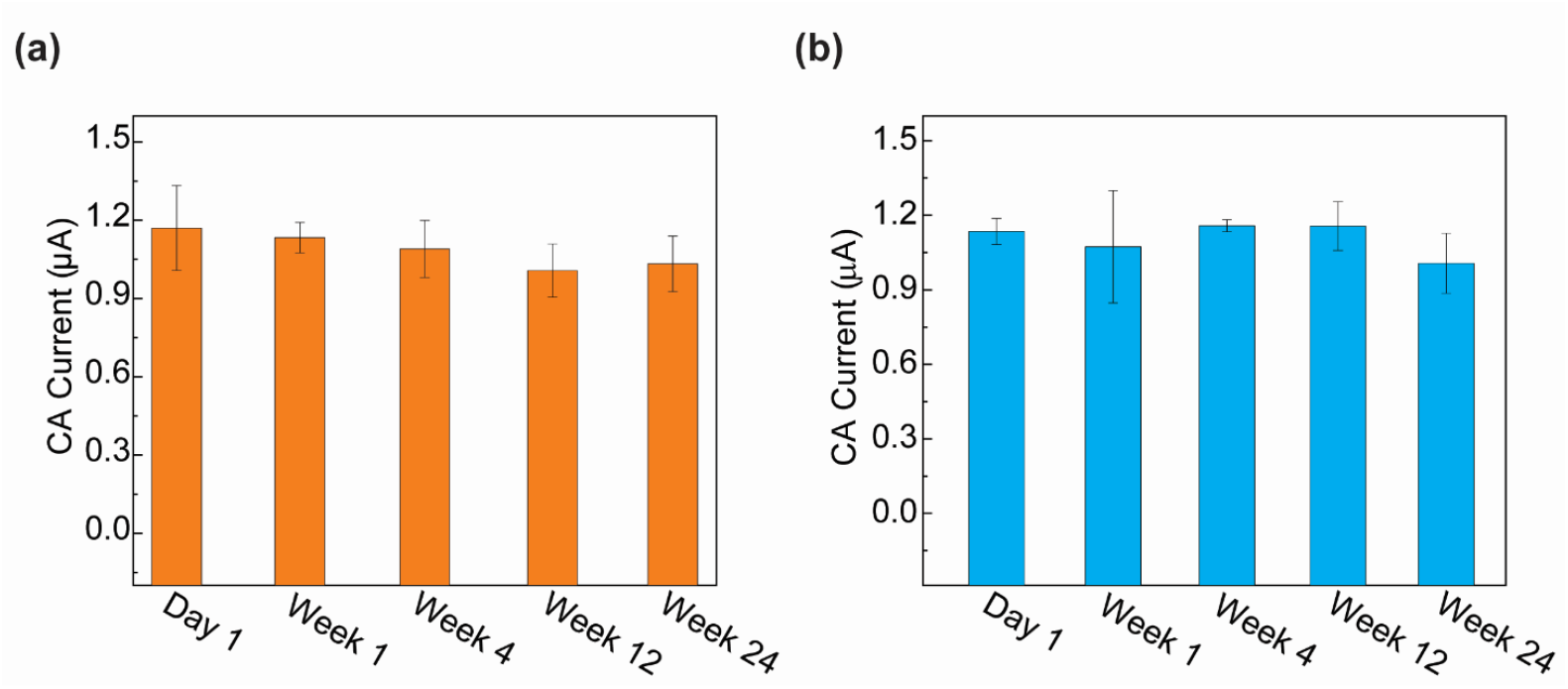
Long-term storage testing of the immunosensor at (a) room temperature and (b) 4 °C in a period of 24 weeks. No significant difference was found between the Day 1 results and the results at all other time points. PBS samples spiked with 6 μg/mL CR3022 IgG were used, and n=4 for all the testing data.

### 3.5 Patient sample testing

A cohort of 20 PCR-tested positive (CoV2+) patient serum samples and 10 pre-pandemic negative (CoV2−) serum samples were tested using our immunosensor, and the experimental setup is shown in **Figure S2**. The CoV2+ serum samples were collected at different time points (1-35 days; see **Table S3**) after the onset of COVID-19 symptoms. **Figure 6a** and **Figure 6b** show the immunosensor current outputs (values listed in **Table S3**) for detecting IgG and IgM of the 20 CoV2+ and 10 CoV2− samples, respectively. The cut-off values for IgG and IgM detection were determined to be 0.259 µA and 0.258 µA, which is defined as the average plus three times the standard deviation of the current outputs measured from the 10 CoV2− samples (**Figure 6a** and **6b**). Based on the calibration curves for CR3022 IgG and IgM detection (**Figure 3c** and **3f**), the equivalent CR3022 IgG and IgM concentrations in the patient samples were calculated (**Figure 6c** and **Table S3**).

**Figure 6.**
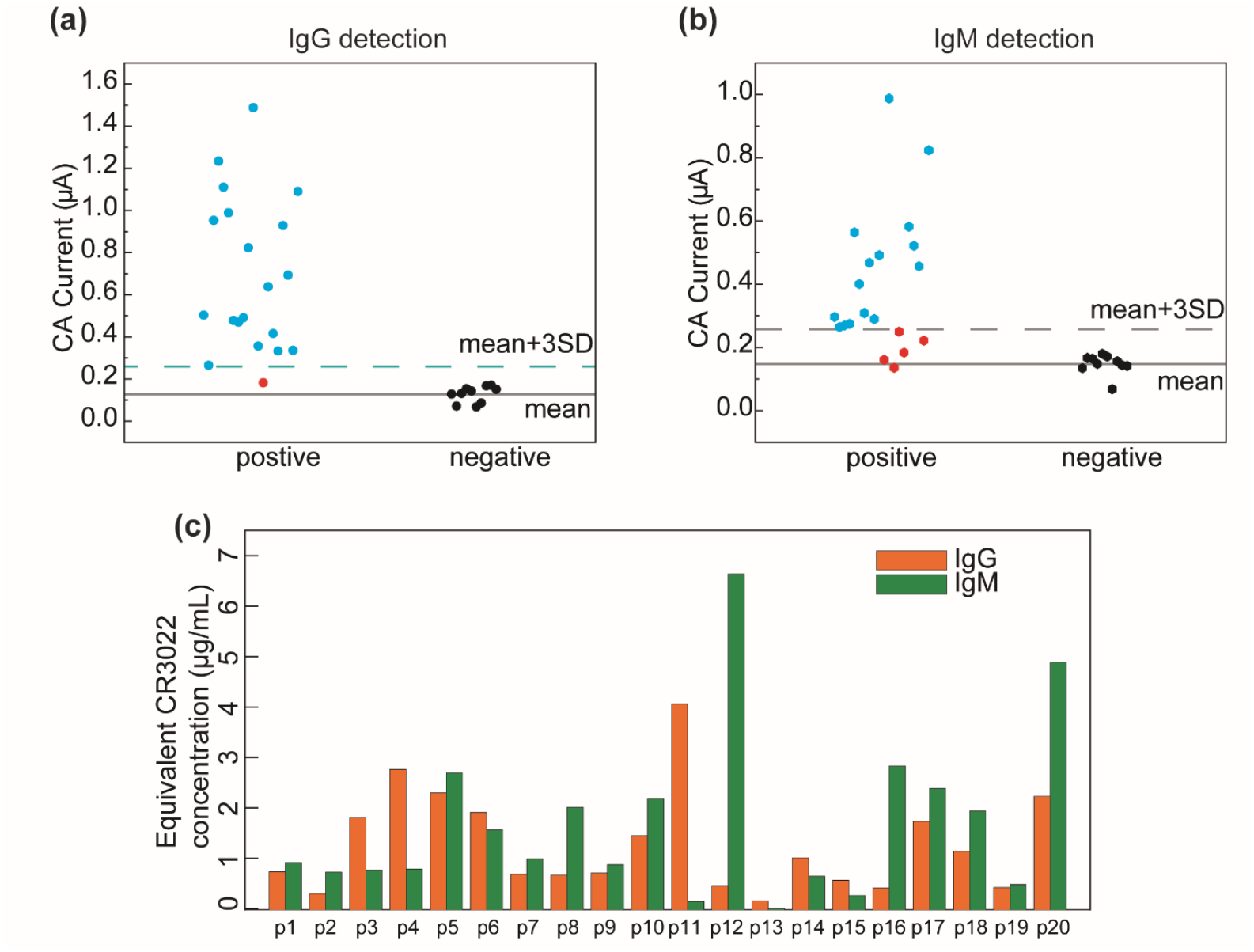
Patient sample testing of the immunosensor. (a)(b) Testing results of (a) IgG and (b) IgM of a cohort of 20 SARS-CoV-2 positive samples and 10 pre-pandemic negative samples. (c) The equivalent CR3022 IgG and IgM concentrations calculated based on the corresponding calibration curves.

From the patient sample testing results, one can see that most of the CoV2+ samples exhibited current signals higher than the cut-off values (IgG+, IgM+) within 1-35 days after symptom onset, indicating the active stage of infection. Only one CoV2+ sample (p13 in **Table S3**) generated both IgG and IgM signals lower than the cut-off values (IgG- and IgM-), which is due to the insufficient immune response of the patient (sample collected on the first day of symptom onset). Detection results corresponding to 30, 33, 33, 34 days after symptom onset in p19, p2, p15, and p11 showed positive IgG and negative IgM levels (IgG+, IgM−), indicating a rapid decay of IgM in these patient samples. Noted that the human immune system of different individuals respond to viral infections dynamically and variably, while the binding activity of all the SARS-CoV-2 anti-S1 protein antibodies could be deemed equivalent to a CR3022 concentration (Ibarrondo et al., 2020). The equivalent concentration can be easily looked up in our calibration curves, and it is always credible and effective for the longitudinal assessment of a single individual. As a result, our immunosensor would serve as a useful tool for immunology studies of COVID-19 including immunity tracking, immunotherapy and immunology diagnosis, where the quantitative evaluation of antibody levels is required.

Due to the limited human sera sample set (20 CoV2+ samples and 10 CoV2− samples) and the non-ideal distribution of the time points of sample collection (16 out of 20 CoV2+ samples were collected during 30-35 days after the symptom onset), we did not calculate sensitivity and specificity of the immunosensor from the current patient sample data. Instead, we compare our SPEEDS platform with other portable biosensors reported in the literature for COVID-19 serology tests, in terms of analytical performance and other technical aspects (**Table 1**). One can see that our SPEEDS platform displays excellent performance in all regards. In particular, our long-term stability facilitates transportation and field application.

**Table 1.** Overviews of SARS-CoV-2 serology analytical devices.

| Biosensor | Analyte and detection method | Analytical performance | Assay time | Portability | Storage stability | Reference |
| --- | --- | --- | --- | --- | --- | --- |
| Chemiluminescent ELISA | Anti-SP IgG<br>Chemiluminescent ELISA | Measurement range: 2–1200 ng/mL<br>LOD: 2 ng/mL | 15 min | CMOS camera used for imaging | Not reported | (Tan et al., 2020a) |
| SARS-CoV-2 RapidPlex | Anti-SP IgG and IgM<br>Graphene-based electrochemical ELISA | Measurement range for IgG and IgM: 50–500 ng/mL<br>LOD: not reported | 13 to 23 min* | Portable system<br>laser-engraved graphene,<br>platinum electrode, | 5 days (at 4 °C) | (Torrente-Rodríguez et al., 2020) |
| Paper-based electrochemical biosensor | Anti-SP IgG and IgM<br>Electrochemical ELISA | Measurement range for IgG and IgM: 1–1000 ng/mL<br>LOD for IgG and IgM : 1 ng/mL | 13 to 63 min* | Portable system,<br>graphene oxide nanosheet<br>modified graphene electrodes,<br>handheld potentiostat and<br>smartphone | 14-day stable storage at 4°C | (Yakoh et al., 2021) |
| Paper-based ELISA | Anti-NP IgG<br>Colorimetric ELISA | Measurement range: 9–50 µg/mL<br>LOD: 9 µg/mL | 15 to 24 min* | Imaging CMOS camera | Not reported | (Kasetsirikul et al., 2020a) |
| Colorimetric lateral flow assay | Anti-NP IgG<br>Nanoparticle-based lateral flow assay | N/A (binary results) | 15 to 20 min | Portable system | Not reported | (Wen et al., 2020) |
| Opto-microfluidic platform | Anti-SP IgG<br>Localized surface plasmon resonance | Measurement range: 0.1 ng/mL–10 µg/mL<br>LOD: 0.08 ng/mL | 30 min | Syringe pump, spectrometer, light source and fiber optics,<br>gold nanospikes | Not reported | (Funari et al., 2020) |
| Vertical flow cellulose-based assay | Anti-NP IgG/IgM (isotype indistinguishable)<br>Colorimetric vertical flow assay | Measurement range for IgG and IgM: 5–40 nM<br>LOD for IgG and IgM: 5 nM | 15 min | Portable system | Not reported | (Kim et al., 2021b) |
| DNA-assisted nanopore sensing | Anti-NP IgG and IgM<br>Electrical resistive pulse DNA nanopore sensing | Measurement range of IgG: 10 ng/mL–100 µg/mL<br>Measurement range of IgM: 50 ng/mL–10 µg/mL<br>LOD of IgG: 10 ng/mL<br>LOD of IgM: 50 ng/mL | ≥40 min * | Patch clamp amplifier, Digidata 1440A data acquisition system | Not reported | (Z. Zhang et al., 2021) |
| Grating-coupled fluorescent plasmonic (GC-FP) biosensor | Anti-SP IgG/IgM/IgA<br>Plasmon-enhanced biosensor | Quantification of antibody titer with serum dilutions ranging from 1:25–1:1600 | 27 min | Fluorescent plasmonic imaging instrument | 4-week stable at room temperature | (Cady et al., 2021) |
| Lateral flow immunoassay immunosensor | Anti-NP IgA<br>Dual lateral flow optical/chemiluminescence immunosensors | Measurement results are in the forms of colorimetric signals (arbitrary units, measured by smartphone reader) and chemiluminescent signals (in relative light unit, measured by CCD camera) | 15 min | Portable system | Not reported | (Roda et al., 2021) |
| Magnetic bead-based Immunoassay | Anti-NP IgG<br>Magnetic bead-based chromogenic ELISA | Log reciprocal dilution:100–500000 | ~ 12 min * | Plate reader | 12-week stable in the fridge | (Huergo et al., 2021) |
| SPEEDS: electrochemical immunosensor | Anti-SP IgG and IgM<br>Quantitative detection by electrochemical ELISA | Measurement range of IgG: 10.1 ng/mL–60 µg/mL<br>Measurement range of IgM: 1.64 ng/mL–50 µg/mL<br>LOD of IgG: 10.1 ng/mL<br>LOD of IgM: 1.64 ng/mL | 13 min | Portable system, handheld potentiostat and smartphone | 24-week stable at room temperature and 4°C | This work |

**Notes:** SP: spike protein, NP: nucleocapsid protein, IgA: immunoglobulin A. \*Estimated assay time based on operation procedure. Accurate time was not reported in the reference.

## 4. Conclusion

We have successfully developed a low-cost and portable SPEEDS platform for the quantitative detection of IgG and IgM antibodies against the SARS-CoV-2 S-protein in human sera. The platform can be used for on-site COVID-19 serological tests and provides accurate results in 13 min. The electrochemical immunosensor can be batch-fabricated at a low material cost and it can be stored at room temperature for at least 24 weeks without performance deterioration. Wide measurement ranges of 10.1 ng/mL − 60 µg/mL and 1.64 ng/mL − 50 µg/mL, have been achieved for IgG and IgM detection, respectively. The immunosensor showed negligible cross-reactivity with non-specific total human IgG and negligible cross-reactivity between CR3022 IgG and CR3022 IgM. A retrospective study on a cohort of 20 COVID-19 positive patient samples and 10 negative samples was conducted to demonstrate the practical use of the SPEEDS platform. We believe that the SPEEDS platform will find important applications in rapid and sensitive serological testing of COVID-19. It can be also extended to other types of serological tests and disease antigen tests.

## Data Availability

All data needed to evaluate the conclusions in the paper are present in the Manuscript and/or the Supplementary Information.

## CRediT Authorship contribution statement

**Ran Peng**: Investigation, Formal analysis, Writing – original draft, Conceptualization, Methodology. **Yueyue Pan**: Investigation, Formal analysis, Writing – original draft, Conceptualization, Methodology. **Zhijie Li**: Investigation. **Zhen Qin**: Investigation. **James M. Rini**: Investigation, Project administration Supervision, Funding acquisition. **Xinyu Liu**: Investigation, Writing, Conceptualization, Supervision, Project administration, Funding acquisition.

## Acknowledgments

This research was supported by the Toronto COVID-19 Action Initiative Program (grant number: 508801), the National Sciences and Engineering Council of Canada (grant numbers: RGPIN-2017-06374 and RGPAS-2017-507980), the Canada Foundation for Innovation (grant number: 30316), and the University of Toronto (through the Percy Edward Hart Professorship to Xinyu Liu).

