## Supplementary Information for "SPEEDS: A Portable Serological Testing Platform for Rapid Electrochemical Detection of SARS-CoV-2 Antibodies"

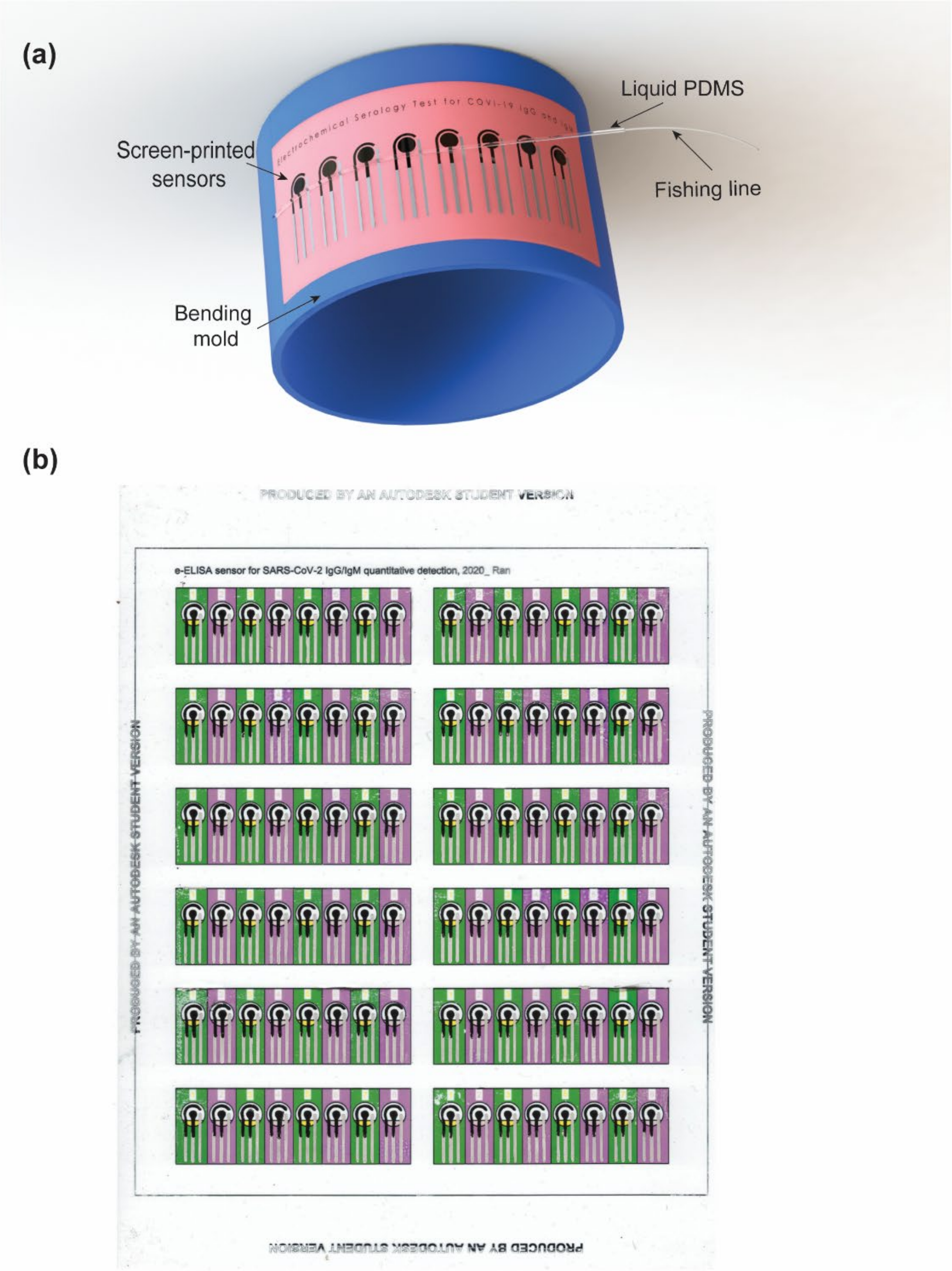


**Fig. S1.** Fabrication of the three-electrode electrochemical immunosensor. (a) A 3D model showing the process of PDMS barrier patterning. The reaction zone was defined by a wax-printed cycle and a thin PDMS line barrier was printed at the intersection of the circular WE zone and its 'tail' connection. (b) A photograph of 96 immunosensors fabricated on a letter-sized PET film.

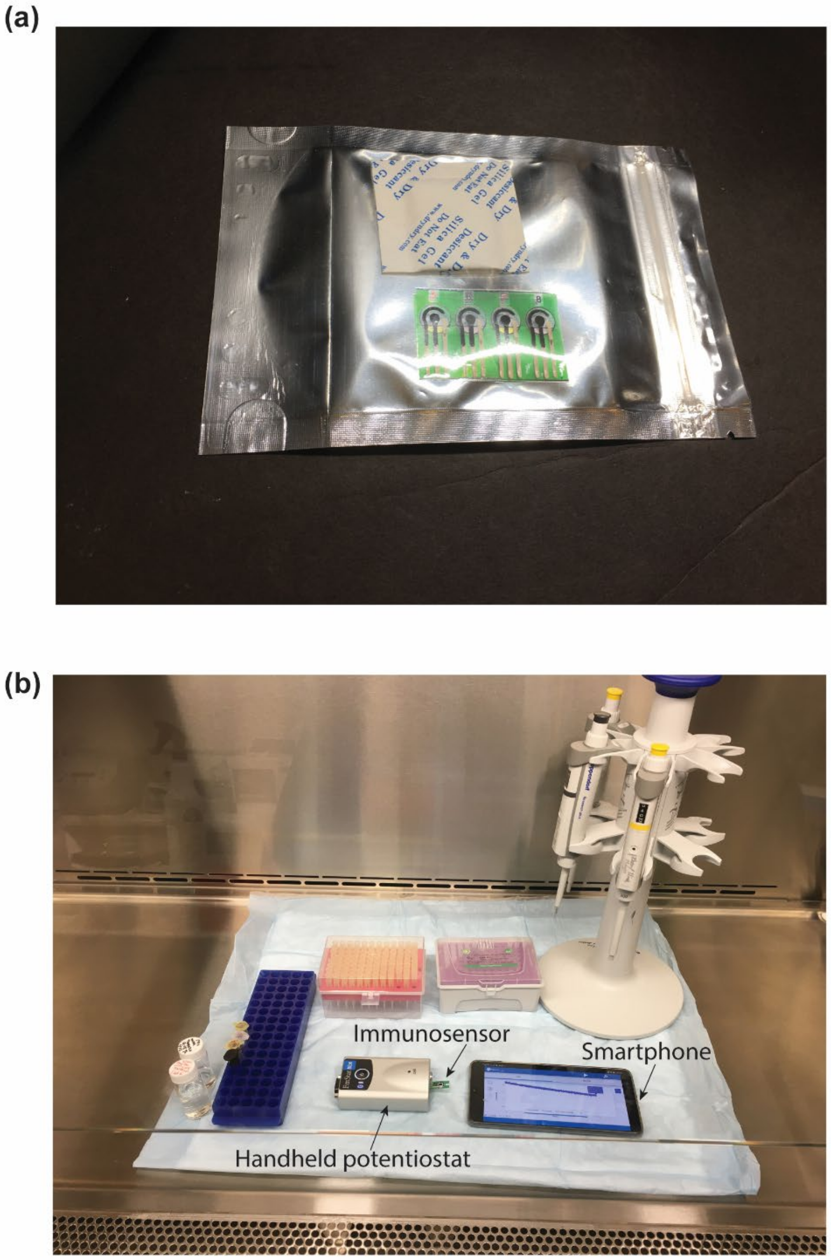


**Fig. S2.** (a) Four immunosensors stored in a nitrogen-infused airtight bag with desiccant inside. (b) The experimental setup for clinical sample testing inside a biosafety cabinet.

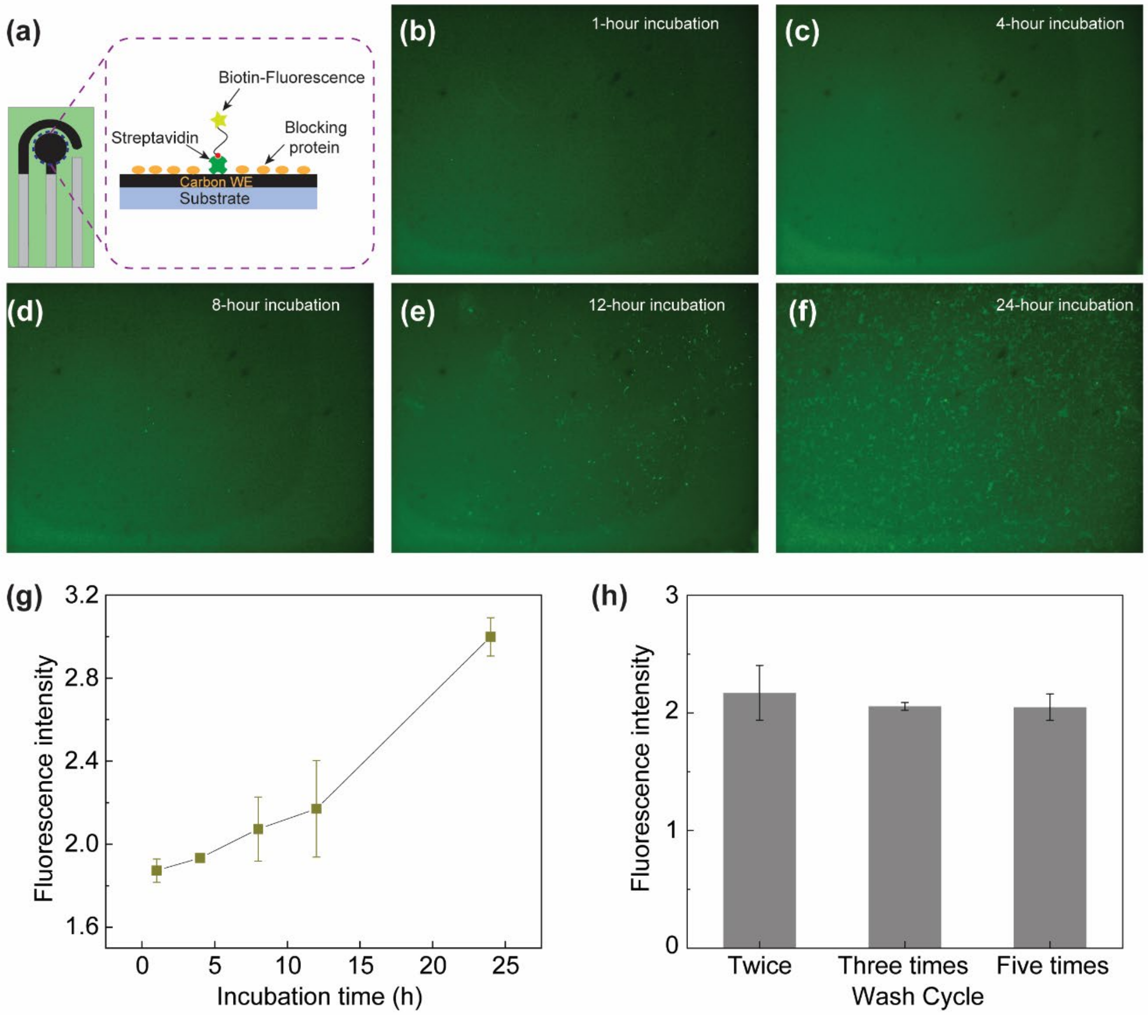


**Fig. S3.** Experimental determination of the incubation and washing conditions for SA immobilization on the WE. (a) The schematic of using biotinylated 5-fluorescein for testing the SA immobilization efficiency. The SA was absorbed on the WE and blocked by blocking proteins, and the fluorescein was captured by the SA and examined by a fluorescence microscope. (b-f) The fluorescence images of WEs, incubated with SA for (b) 1 h, (c) 4 h, (d) 8 h, (e) 12 h, and (f) 24 h, then labelled with biotin-5-fluorescein conjugate, and finally washed with PBS for three times. (g) Average fluorescence intensity of the WEs after SA incubation for different durations (n = 4). (h) The fluorescence intensity of the WEs after SA incubation of 12 h, treated with biotin-5-fluorescein conjugate, and washed for twice, three and five times (n=4).

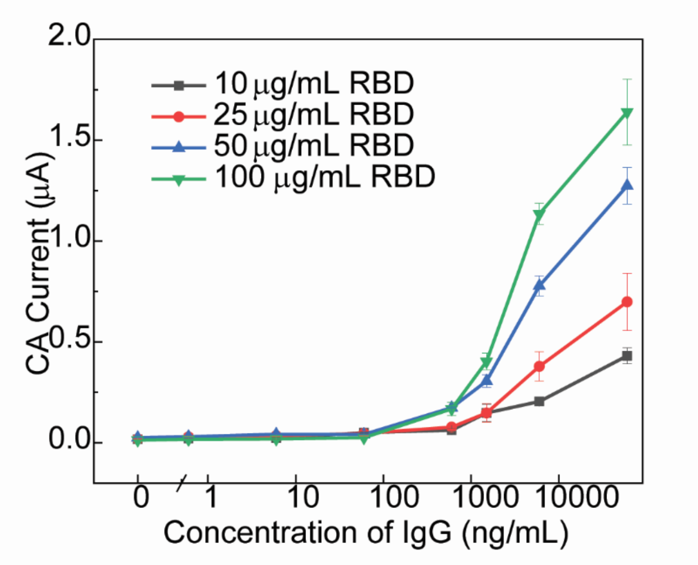


**Fig. S4.** Calibration curves of CR3022 IgG detection using spiked PBS on immunosensors prepared with four different concentrations of the biotinylated RBD capture probe (n=4). The variations of the CA current from different concentrations of RBD can be seen in the high concentration range (> 600 ng/mL).

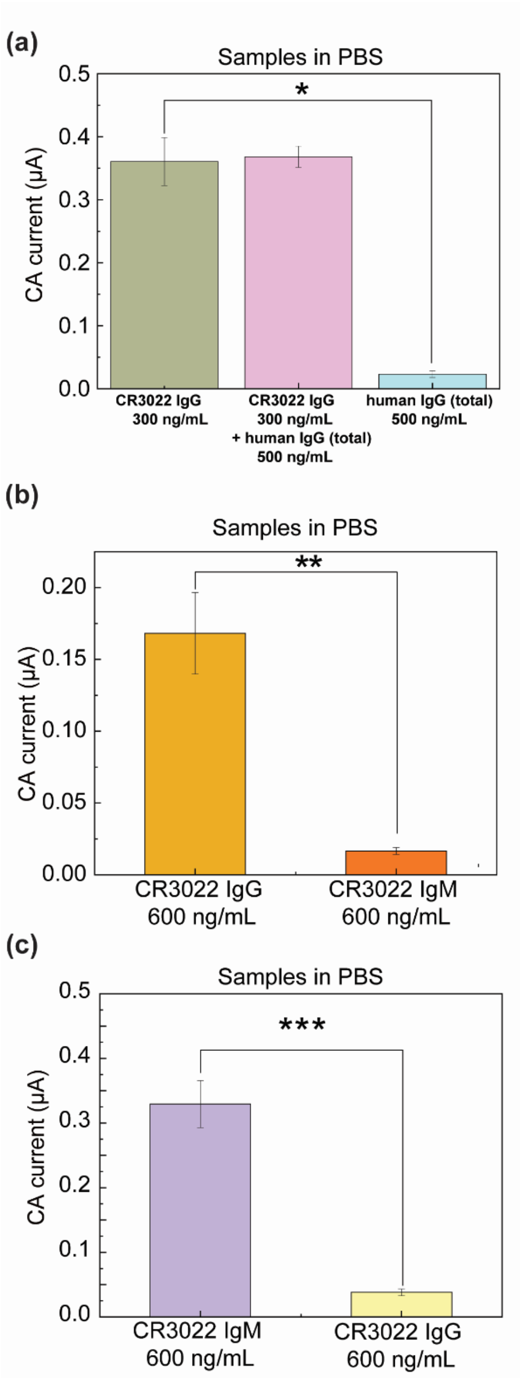


**Fig. S5.** The cross-reaction testing of anti-SARS-CoV-2 IgG and IgM detection in PBS (n=5). (a) The cross-reactivity data of CR3022 IgG detection using total human IgG as the interference protein. *$p=4.72\times10^{-8}$. (b) The cross-reactivity data of CR3022 IgG detection using CR3022 IgM as the interference protein. **p-value=$2.29\times10^{-6}$; (c) The cross-reactivity data of CR3022 IgM detection using CR3022 IgG as the interference protein. ***p-value=$1.06\times10^{-7}$.

**Table S1.** The cost breakdown of one test.

| Material | Cost for a single test (USD) |
|---|---|
| **PET film** | 0.00718 |
| **Protection film** | 0.00124 |
| **Airtight bags** | 0.0259 |
| **Desiccants** | 0.00797 |
| **Carbon ink** | 0.00957 |
| **Ag/AgCl ink** | 0.120 |
| **PDMS** | 0.0000167 |
| **$H_2SO_4$** | 0.000360 |
| **Streptavidin** | 0.391 |
| **Biotinylated RBD** | 1.50 |
| **Blocking reagent for ELISA** | 0.00219 |
| **Immunoassay stabilizer** | 0.0174 |
| **ALP-conjugated goat anti-human IgG/ ALP-conjugated goat anti-human IgM** | 0.0171 |
| **pAPP** | 0.00128 |
| **Total cost** | 2.10 |

**Table S2** Fitting parameters. The calibration curves under two detection modes are fitted with the Boltzmann model: $y = f(x) = \frac{A_1 - A_2}{1 + e^{(x - x_0)/dx}} + A_2$, where $y$ is the CA current in amperes (A) and $x$ is the concentration of antibody in ng/mL.

| | $A_1$ | $A_2$ | $x_0$ | $dx$ | $R^2$ |
|---|---|---|---|---|---|
| IgG calibration curve | $-1.20\times 10^{-4}$ | $1.91\times 10^{-6}$ | $-1.15 \times 10^{4}$ | $2.75 \times 10^{3}$ | 0.981 |
| IgM calibration curve | $-2.79\times 10^{-4}$ | $1.68\times 10^{-6}$ | $-4.29 \times 10^{4}$ | $8.25 \times 10^{3}$ | 0.965 |

**Table S3** Testing results of five-fold-diluted human serum samples.

| Positive patient samples (CoV2+) | Days since symptom onset | Average IgG test CA current (μA) | IgG concentration (ng/mL) | Average IgM test CA current (μA) | IgM concentration (ng/mL) |
|---|---|---|---|---|---|
| p1 | 33 | 0.503 | 731 | 0.297 | 915 |
| p2 | 33 | 0.266 | 297 | 0.265 | 726 |
| p3 | 33 | 0.953 | 1800 | 0.27 | 758 |
| p4 | 33 | 1.23 | 2760 | 0.275 | 786 |
| p5 | 34 | 1.11 | 2300 | 0.564 | 2690 |
| p6 | 34 | 0.99 | 1910 | 0.401 | 1560 |
| p7 | 34 | 0.479 | 683 | 0.309 | 987 |
| p8 | 33 | 0.47 | 666 | 0.468 | 2010 |
| p9 | 34 | 0.491 | 707 | 0.29 | 876 |
| p10 | 34 | 0.823 | 1450 | 0.492 | 2180 |
| p11 | 34 | 1.49 | 4060 | 0.161 | 141 |
| p12 | 8 | 0.357 | 455 | 0.988 | 6640 |
| p13 | 1 | 0.182 | 159 | 0.136 | 4.41 |
| p14 | 4 | 0.638 | 1010 | 0.250 | 641 |
| p15 | 33 | 0.416 | 564 | 0.184 | 265 |
| p16 | 33 | 0.333 | 414 | 0.582 | 2830 |
| p17 | 33 | 0.929 | 1730 | 0.522 | 2380 |
| p18 | 9 | 0.694 | 1130 | 0.457 | 1940 |
| p19 | 30 | 0.336 | 419 | 0.222 | 479 |
| p20 | 35 | 1.09 | 2230 | 0.824 | 4890 |
| Negative patient samples (CoV2−) | | | | | |
| n1 | N/A | 0.129 | N/A | 0.135 | N/A |
| n2 | N/A | 0.0715 | N/A | 0.167 | N/A |
| n3 | N/A | 0.132 | N/A | 0.165 | N/A |
| n4 | N/A | 0.154 | N/A | 0.148 | N/A |
| n5 | N/A | 0.144 | N/A | 0.180 | N/A |
| n6 | N/A | 0.0683 | N/A | 0.171 | N/A |
| n7 | N/A | 0.0865 | N/A | 0.068 | N/A |
| n8 | N/A | 0.168 | N/A | 0.157 | N/A |
| n9 | N/A | 0.171 | N/A | 0.144 | N/A |
| n10 | N/A | 0.152 | N/A | 0.142 | N/A |

.
